# Too much to mask: determinants of sustained adherence to COVID-19 preventive measures among older Syrian refugees in Lebanon

**DOI:** 10.1101/2022.05.10.22274919

**Authors:** Nisreen Salti, Stephen J. McCall, Berthe Abi Zeid, Noura El Salibi, Marwan Alawieh, Zeinab Ramadan, Hala Ghattas, Sawsan Abdulrahim

## Abstract

Lebanon has battled the COVID-19 pandemic in the midst of an economic crisis. The evolution of the pandemic and a fragile health system have meant that public health policy has had to rely heavily on non-pharmaceutical interventions for disease control. However, changes in disease dynamics and pandemic fatigue have meant that disease control policies need to be updated. Identifying variables associated with adherence to non-pharmaceutical preventive practices, particularly for vulnerable groups, can therefore help inform and refine interventions in the face of pandemic fatigue and changing disease dynamics. Using recent and timely data on older (50 years and above) Syrian refugees in Lebanon, this paper explores the determinants of adherence to two non-pharmaceutical COVID-19 prevention measures (wearing a mask and avoiding social gatherings) among this high-risk subgroup in a vulnerable population. Among respondents who report adhering to these measures, the paper also identifies the determinants of sustained adherence over a period of 6 months. The findings suggest that older refugees and those less educated are less likely to wear a mask, and refugees living in informal tented settlements are more likely to relent on preventive practices within 6 months. Individuals with chronic diseases who initially report avoiding social gatherings are also likelier to desist than those without chronic illness.

The lower continued adherence to mask wearing among residents of informal tented settlements points to factors beyond pandemic fatigue and that should be taken into consideration in devising measures for disease control: the potential for community-based norms to determine individual-level behavior. Recognizing the pivotal effect of community-based norms in settings such as informal tented settlements is essential in adapting current policy and designing future interventions.

## I. Background and motivation

Lebanon is suffering a confluence of crises: an economic and financial collapse that started in 2019 and has slashed Gross Domestic Product (GDP), doubled the poverty rate, and mired the country in hyperinflation; the COVID-19 pandemic since early 2020; and the explosion at the Port of Beirut in August of 2020 that caused extensive human and material damage. As a result, the country’s healthcare system has been pushed to the brink of collapse (Isma’eel et al., 2020; MSF, 2021; Shallal et al., 2021).

The emergence of new variants of the coronavirus, the ensuing surges in recorded cases, as well as the slow rollout of vaccination campaigns have meant that Lebanon, like many other countries, continues to rely on non-pharmaceutical interventions (NPIs) to curb the spread of the COVID-19 pandemic, until herd immunity is reached. In fact, with Lebanon’s economy already in freefall and its public health infrastructure severely compromised, low cost NPIs can play a pivotal role in preventing and controlling the spread of the virus. Research suggests that mask wearing and reducing contact with others are effective NPIs for preventing infection from COVID-19 (Cheng et al., 2020; Siedner et al., 2020; Teslya et al., 2020; Xiao & Torok, 2020). However, with the pandemic about to enter its third year, continued adherence to some of these preventive measures, such as staying at home except for essential outings, restricting travel, and limiting visitors, is proving increasingly difficult to maintain. Indeed, ‘pandemic fatigue’ has meant that even adherence to far less costly preventive measures, such as mask wearing, is on the wane.

Continued reliance on NPIs implies that the potential effects of ‘pandemic fatigue’ on adherence should be of concern (WHO, 2020), all the more so in a context like Lebanon in which an economic crisis has made daily life increasingly challenging and has largely depleted people’s motivation to comply with new behavioral recommendations. Among the most exposed to both the economic crisis and the public health emergency in Lebanon are Syrian refugees. Beyond their socio-economic vulnerability even prior to the economic crisis (Chaaban et al., 2020; UNHCR et al., 2019), Syrian refugees are also more vulnerable to COVID-19 because they tend to live in crowded conditions with limited access to sanitation and public health infrastructure (Fouad et al., 2021). Among Syrian refugees, older adults are particularly exposed to the risk of severe morbidity and death from COVID-19.

This paper uses recent and timely data on older Syrian refugees (50+) to explore the determinants of adherence to wearing a mask and avoiding social gatherings (such as weddings and funerals) among this high-risk subgroup in a vulnerable population, as well as the determinants of sustained adherence over a period of 6 months among those who report to abide by these recommendations. The focus is on these two practices since they are among the lowest cost, effective COVID-19 prevention NPIs.

In the face of the enormous economic and logistical challenges faced by public health policy makers in Lebanon today, identifying the factors associated with adherence to low-cost and effective preventive measures, and factors associated with waning adoption of these measures by those most at risk among vulnerable populations may go a long way in informing policies aimed at curbing the spread of COVID-19, averting placing further pressure on an already frail public health infrastructure, and fine-tuning messaging for interventions such as mobility restrictions and vaccination in the face of pandemic fatigue.

## II. Literature review

A growing literature has emerged on the correlates and factors associated with the use of masks and the adoption of other preventive measures globally (Adjodah et al., 2021; Badillo-Goicoechea et al., 2021). The literature is also rich with country studies examining the predictors and correlates of mask wearing and other preventive measures. Daoust (2020) identified the correlates of compliance with preventive measures across 27 countries, with a focus on older adults. In the context of Lebanon, some work has tried to identify correlates of adherence to preventive measures (Domiati et al., 2020), sometimes for some subpopulations (Abou-Abbas et al., 2020; Nasser et al., 2020; Sakr et al., 2021), while other work has looked at the role of specific interventions in affecting or mediating adherence (Melki et al., 2020).

While the identification of such correlates may have been informative for policy makers trying to devise prevention and control policies, COVID-19 control strategies have to reckon with the fact that people’s behavior also changes over time. Changing behavior means that subgroups that were initially adherent may not remain so for long, so pandemic response policies need to be updated accordingly. However, research that aims to shed light on the dynamics of adherence over time is more scant.

Chan et al. (2021) use repeated cross-sectional data to examine the socio-economic and demographic correlates of changing adherence to social distancing and personal hygiene measures in Hong Kong during the first and third waves of COVID-19. They find improved adherence to mask wearing and decreasing compliance with social distancing measures (including avoidance of gatherings, of public places, and of international travel).

This is in line with evidence from multi-country studies which suggests that adherence to sensitizing behaviors such as physical distancing has decreased, while adherence to habituating behaviors such as mask wearing has increased (Petherick et al., 2021). Like Chan et al. (2021), Petherick et al. (2021) also base their results on observational data, but their repeated cross sections are more frequent and span 14 countries.

Unlike both Chan et al. (2021) and Petherick et al (2021), we do not seek to measure cross-population changes in compliance over time. Instead, we focus on one subgroup (older adults) in one subpopulation (Syrian refugees), follow the same individuals over time, and track their self-reported compliance behavior on two occasions that are 6 months apart. The timing of our sample means that unlike Chan et al. (2021), we are not limited to observations only during periods of surges of COVID-19 cases.

Research on the dynamics of compliance in Lebanon is still very preliminary: Makki et al. (2020) document some evidence in support of pandemic fatigue in Lebanon, but their findings are based on a very small sample (n=30, only some of which are from Lebanon) from a pilot study. With their sample size, they are unable to identify the correlates of behavioral fatigue in adherence to preventive measures.

This paper shares some of the aims of Daoust (2020), but rather than only profiling compliers by using cross-sectional data to identifying correlates of adherence to a preventive measure, we also attempt to identify individual-level correlates of compliers who maintain compliance over 6 months vs. those who relent on it. If uncovering the correlates of adherence to preventive measures was informative for COVID-19 response policies early on during the pandemic, identifying the characteristics associated with waning compliance is of relevance to updating these policies, fine-tuning the target populations, and sharpening the messaging in response to ‘pandemic fatigue’.

## III. Data description and methods

### A. Data and methods

The analysis is based on a sample of 3,839 Syrian refugees 50 years of age or older residing in Lebanon, drawn from households with at least one adult known to be 50 years or older from a list of beneficiary households of a humanitarian organization. We use data from two phone surveys conducted with participants six months apart, the first between September and December of 2020, and the second between February and May 2021. The surveys include information on demographic characteristics of the individual and their household, ownership of assets, labor market status, health status, the receipt of cash assistance, including COVID-19 related cash assistance, COVID-related behaviors, food and water insecurity, decision making and social support, exposure to violence and security risks.

Respondents were asked on 2 occasions 6 months apart about their adherence to two different practices related to recommended preventive measures. The recommended practices are: (i) wearing a mask and (ii) avoiding social occasions (such as weddings and funerals). The first is a habituating behavior, the second a sensitizing one, but they are both associated with relatively lower costs on the complier when compared to the other COVID-19 control recommendations such as sheltering in place and avoiding travel.

Summary statistics are presented in Table 1. Individual level characteristics include demographic information (sex, age, marital status), education, labor market status, an indicator of whether the COVID-19 preventive behaviors are the respondent’s own decision, smoker status, and an indicator for the respondent having a chronic health condition. Adherence variables include wearing a mask and avoiding social gatherings. Household-level characteristics include household size, assets, an indicator for severe food insecurity, an indicator for receiving cash assistance, an indicator for living in an ITS, in addition to district of residence and month of data collection.

**Table 1:**
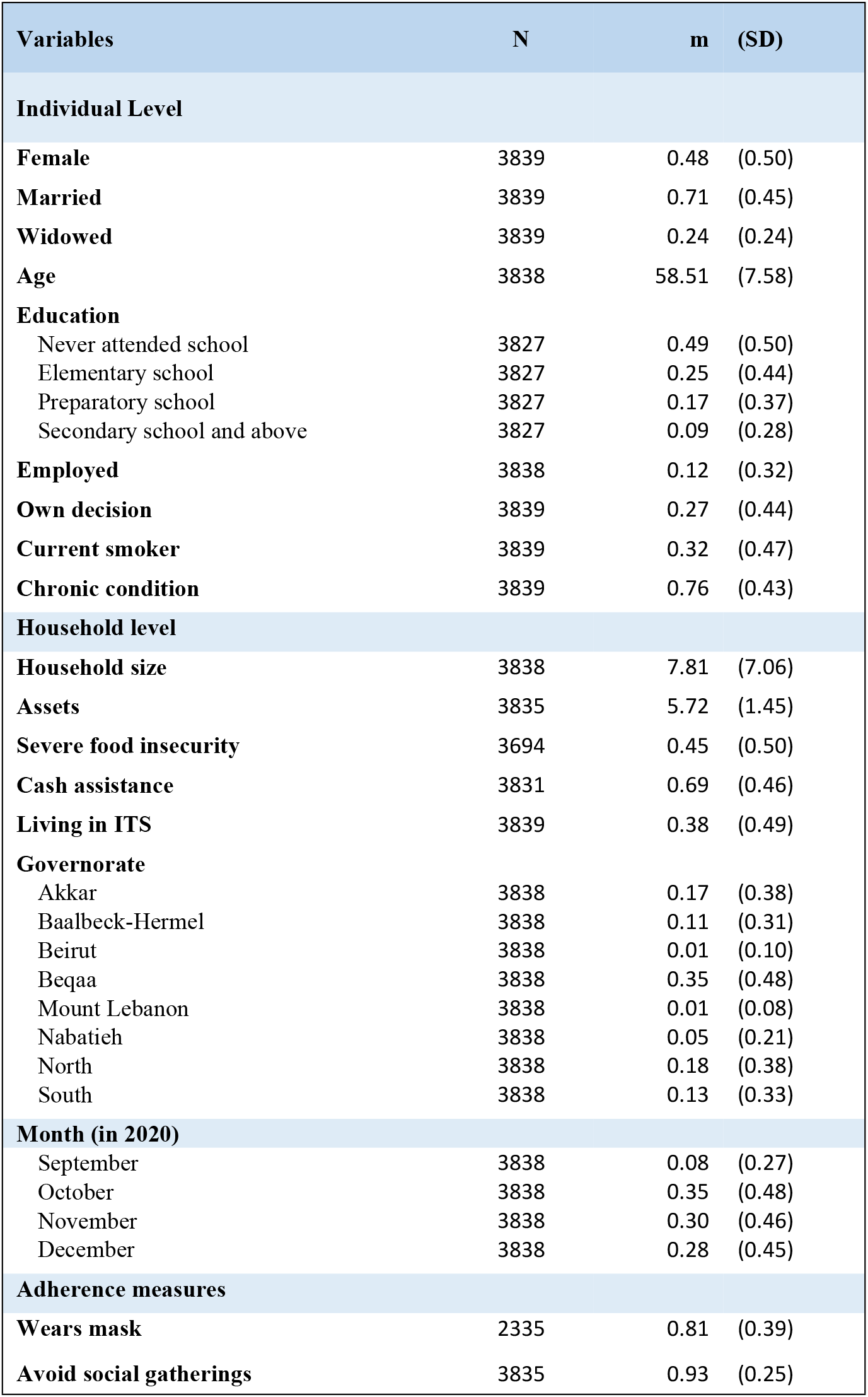
Summary statistics.

Linear probability models are used to determine the correlates of adherence to each of the two preventive behaviors separately using the entire sample, and determinants of sustained adherence 6 months later among those who initially report adherence. Linear probability models use the discrete adherence behavior variables as dependent variables and investigate partial correlations with some of the associated socio-demographic, labor market, health and household level variables. Regressions of adherence have individual-level and household-level regressors. Individual-level variables include socio-demographic characteristics (sex, age, marital status, household size, education), labor market status (employment status), health (smoker status, chronic conditions), self-reported susceptibility to COVID-19, a variable measuring the decision-making process in the household about adherence to preventive behaviors. Household-level variables include socio-economic variables (food security status as measured using the Food Insecurity Experience Scale, asset ownership^1^), governorate of residence, an indicator of residence in an informal tented settlement (ITS), and an indicator measuring receipt of COVID-19 cash assistance. Regressions of sustained adherence are run on the subsample of initial adherents.

Table 2 reports summary statistics for respondents who initially adhere to mask wearing (Panel A), and for respondents who initially adhere to avoiding social gatherings (Panel B). These also include variables measuring the respondent’s and the household’s COVID-19 history, changes in their labor market status and earnings, and measures of respondents moving, including moving into or out of ITSs.

**Table 2:** Summary statistics for subsample of adherents.

Panel A: Initial adherents to mask wearing
| Variables | N | m | (SD) |
| --- | --- | --- | --- |
| **Sustained mask wearing** | 1452 | 0.36 | (0.48) |
| **Female** | 1452 | 0.46 | (0.50) |
| **Married** | 1452 | 0.74 | (0.43) |
| **Widowed** | 1452 | 0.21 | (0.41) |
| **Age** | 1452 | 57.86 | (7.22) |
| **Education** |  |  |  |
| Never attended school | 1449 | 0.49 | (0.50) |
| Elementary school | 1449 | 0.26 | (0.44) |
| Preparatory school | 1449 | 0.16 | (0.37) |
| Secondary school and above | 1449 | 0.08 | (0.27) |
| **Employed** | 1452 | 0.07 | (0.25) |
| **Job loss (due to COVID-19)** | 1452 | 0.16 | (0.36) |
| **Salary reduction (due to COVID-19)** | 1452 | 0.16 | (0.37) |
| **Own decision** | 1452 | 0.33 | (0.47) |
| **Current smoker** | 1452 | 0.40 | (0.49) |
| **Chronic condition** | 1452 | 0.72 | (0.45) |
| **Had COVID-19** | 1452 | 0.02 | (0.15) |
| **Household level** |  |  |  |
| **Household COVID-19** | 1452 | 0.01 | (0.05) |
| **Household size** | 1452 | 7.82 | (4.02) |
| **Assets** | 1452 | 5.59 | (1.48) |
| **Severe food insecurity** | 1425 | 0.40 | (0.49) |

<sup>1</sup> Asset ownership is a variable counting the number of affirmative answers to questions about the ownership of the following 15 assets: car, motorbike/scooter, van/pickup truck, bicycle, gas stove, oven, refrigerator, iron, heater, water heater, washing machine, TV, computer, mobile phone, access to the internet.

|  |  |  |  |
| --- | --- | --- | --- |
| **Cash assistance** | 1449 | 0.79 | (0.41) |
| **Living in ITS** | 1452 | 0.56 | (0.50) |
| **Moved to ITS** | 1452 | 0.13 | (0.33) |
| **Moved from ITS** | 1452 | 0.13 | (0.33) |
| **Governorate** |  |  |  |
| Akkar | 1452 | 0.15 | (0.36) |
| Baalbeck-Hermel | 1452 | 0.14 | (0.35) |
| Beirut | 1452 | 0.01 | (0.07) |
| Beqaa | 1452 | 0.41 | (0.49) |
| Mount Lebanon | 1452 | 0.01 | (0.08) |
| Nabatieh | 1452 | 0.04 | (0.19) |
| North | 1452 | 0.14 | (0.35) |
| South | 1452 | 0.10 | (0.30) |
| **Month (in 2021)** |  |  |  |
| February | 1452 | 0.12 | (0.33) |
| March | 1452 | 0.52 | (0.50) |
| April | 1452 | 0.16 | (0.37) |
| May | 1452 | 0.20 | (0.40) |

Panel B: Initial adherents to avoiding social gatherings
| **Variables** | **N** | **m** | **(SD)** |
| --- | --- | --- | --- |
| **Sustained avoiding social gatherings** | 2789 | 0.87 | (0.33) |
| **Female** | 2789 | 0.48 | (0.50) |
| **Married** | 2789 | 0.71 | (0.45) |
| **Widowed** | 2789 | 0.25 | (0.43) |
| **Age** | 2789 | 58.42 | (7.58) |
| **Education** |  |  |  |
| Never attended school | 2783 | 0.49 | (0.50) |
| Elementary school | 2783 | 0.26 | (0.44) |
| Preparatory school | 2783 | 0.16 | (0.37) |
| Secondary school and above | 2783 | 0.08 | (0.28) |
| **Employed** | 2789 | 0.07 | (0.25) |
| **Job loss (due to COVID-19)** | 2789 | 0.13 | (0.34) |
| **Salary reduction (due to COVID-19)** | 2789 | 0.14 | (0.34) |
| **Own decision** | 2789 | 0.30 | (0.46) |
| **Current smoker** | 2789 | 0.37 | (0.48) |
| **Chronic condition** | 2789 | 0.70 | (0.46) |
| **Had COVID-19** | 2789 | 0.04 | (0.20) |
| **Household level** |  |  |  |
| **Household COVID-19** | 2789 | 0.01 | (0.07) |
| **Household size** | 2789 | 7.85 | (4.08) |
| **Assets** | 2786 | 5.73 | (1.45) |
| **Severe food insecurity** | 2743 | 0.40 | (0.49) |
| **Cash assistance** | 2783 | 0.78 | (0.41) |
| **Living in ITS** | 2789 | 0.38 | (0.48) |
| **Moved to ITS** | 2789 | 0.11 | (0.31) |
| **Moved from ITS** | 2789 | 0.10 | (0.30) |
| **Governorate** |  |  |  |
| Akkar | 2789 | 0.18 | (0.39) |
| Baalbeck-Hermel | 2789 | 0.11 | (0.31) |
| Beirut | 2789 | 0.01 | (0.08) |
| Beqaa | 2789 | 0.35 | (0.48) |
| Mount Lebanon | 2789 | 0.01 | (0.08) |
| Nabatieh | 2789 | 0.04 | (0.21) |
| North | 2789 | 0.17 | (0.38) |
| South | 2789 | 0.13 | (0.33) |
| **Month (in 2021)** |  |  |  |
| February | 2789 | 0.07 | (0.26) |
| March | 2789 | 0.32 | (0.47) |
| April | 2789 | 0.27 | (0.45) |
| May | 2789 | 0.33 | (0.47) |

## IV. Results and discussion

### A. Mask wearing and avoiding large social gatherings

Table 3 shows the results of the linear probability model of wearing a mask (column 1-2) and avoiding social gatherings (column 3-4).

**Table 3:**
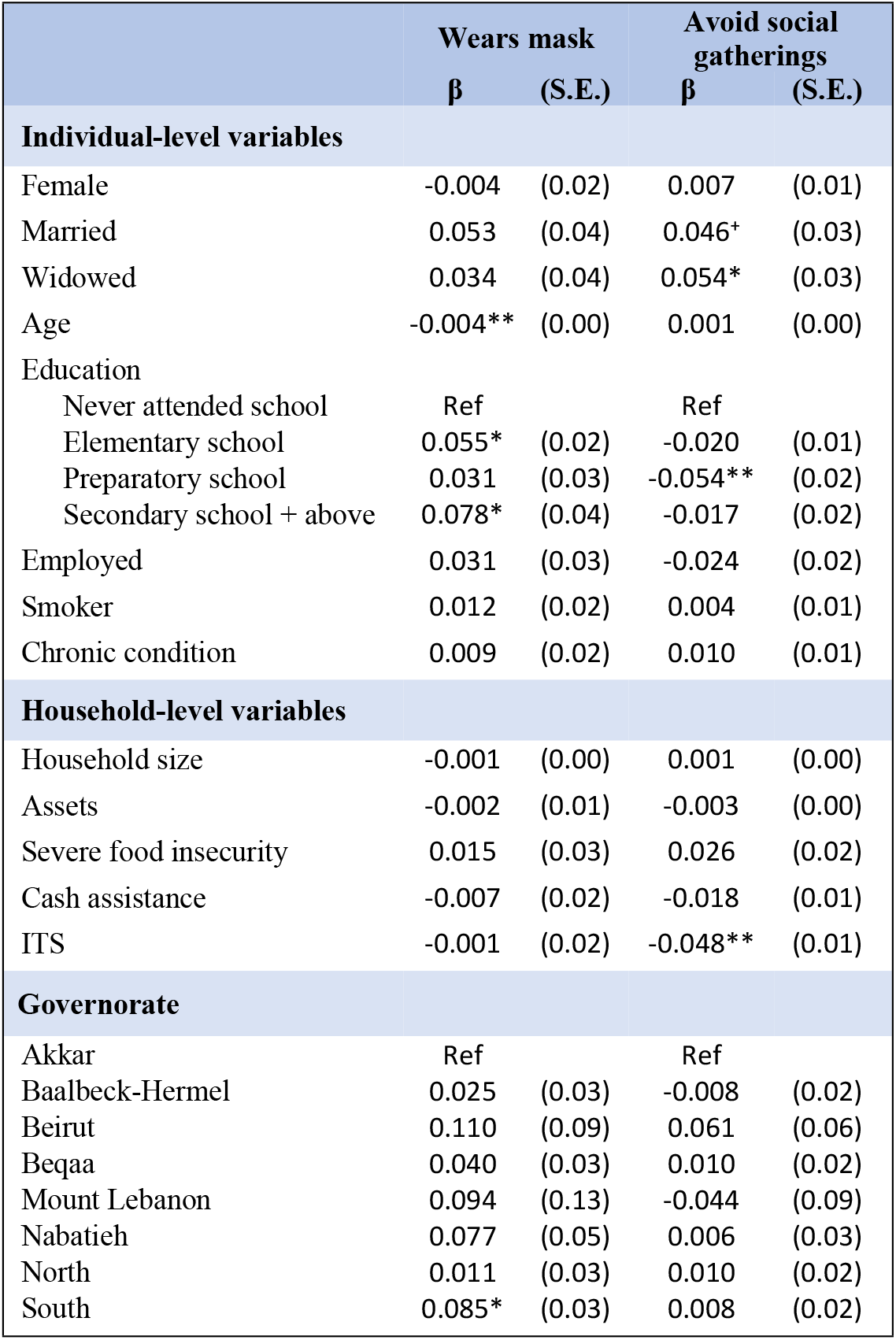
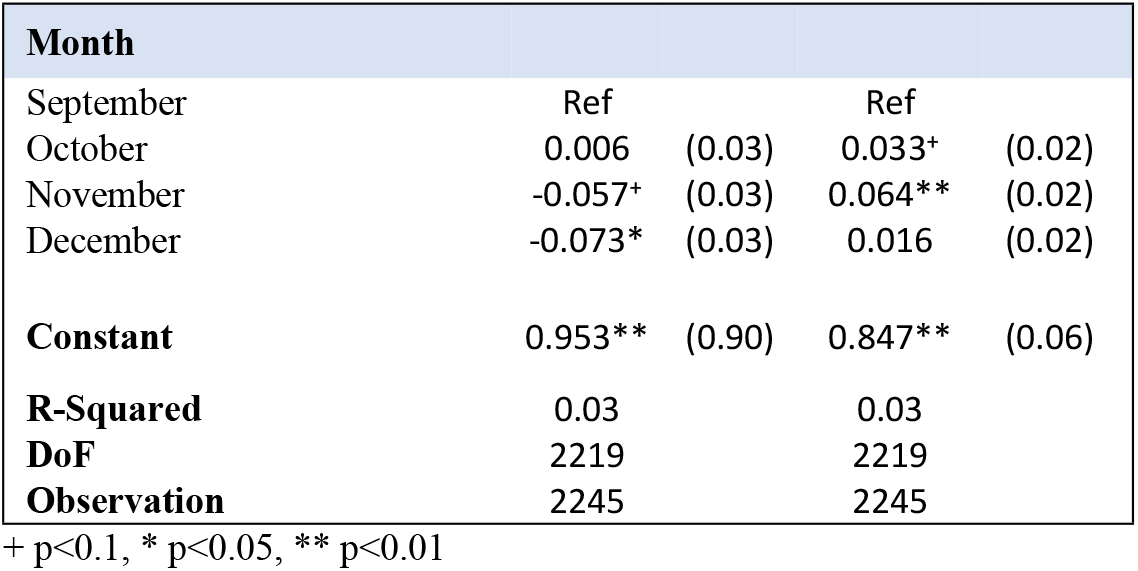
Adherence to preventive measures.

Although the literature reports a gender difference in mask wearing (Cassino & Besen-Cassino, 2020; Chuang & Liu, 2020; Haischer et al., 2020; Okten et al., 2020), we find no difference between males and females in mask wearing among older Syrian refugees.

Whereas previous studies have shown that over the entire age range of the adult population, mask wearing tends to increase with age (Badillo-Goicoechea et al., 2021; Haischer et al., 2020), we find that among elderly Syrian refugees, the older are significantly less likely to wear a mask, with the probability of wearing a mask 2 percentage points lower for every 5 additional years of age.

Education is associated with higher likelihood of wearing a mask, with respondents with elementary and secondary or higher education significantly more likely to wear a mask than respondents with no schooling (by 5.5 percentage points and 7.8 percentage points respectively). This is in line with findings from the literature (Badillo-Goicoechea et al., 2021) (Sileshi, 2021) (Ditekemena, 2021). No other individual-level correlate is found to be significantly associated with mask wearing.

We find a geographic difference in the likelihood of wearing a mask with residents of the governorate of the South significantly more likely to mask than residents of Akkar in the north of the country. This is consistent with findings from an earlier nationally representative survey that finds lower levels of knowledge and practice on COVID-19 prevention in Akkar and the North even among Lebanese residents (UNICEF, 2020).

The only other significant correlate of mask wearing is a time fixed effect: there is a decline in the likelihood of masking over time, with respondents in November and December significantly less likely to report using masks than respondents in September and October of 2020, likely indicating generalized pandemic fatigue.

For self-reported avoidance of social gatherings (columns 3-4), we find no significant differences in gender or age. Married and widowed respondents were slightly likelier to report adhering to this measure than respondents who are never married or separated, which is unsurprising. The other significant individual-level correlate that we find is education, with respondents with an intermediate level of education less likely to abide by the recommendation to avoid social gatherings than respondents with no education by 5.4 percentage points (significant at 5%).

Unlike for mask wearing, we find no geographic differences, but we do find that residents of ITSs are significantly less likely to avoid social gatherings by 4.8 percentage points. This result is unsurprising given the crowdedness and physical proximity of shelters within ITSs, as well as the strict movement restrictions that were imposed early on in the pandemic that largely confined ITS residents to their camps, but didn’t police their movement within the settlement (Moawad & Andres, 2021).

Month fixed effects show that respondents were significantly likelier to avoid social gatherings in October and November than they were in September of 2020. This corresponds with a period of rising daily cases from September onward that culminated in the imposition of a two-week country-wide lockdown in mid-November (Koweyes et al., 2021).

Overall, the explanatory power of the regressions is low: adherence to preventive measures is relatively new behavior based on complex psychological, cognitive and emotional factors to do with perceived risk, discipline, and adaptability. We do not expect to be able to capture much of the complexity of adherence to preventive measures by looking at partial correlations with a small set of measurable characteristics.

### B. Sustained adherence

Results of the linear probability regressions of sustained adherence over six months for wearing a mask and avoiding social gatherings are shown in Table 4.

**Table 4:**
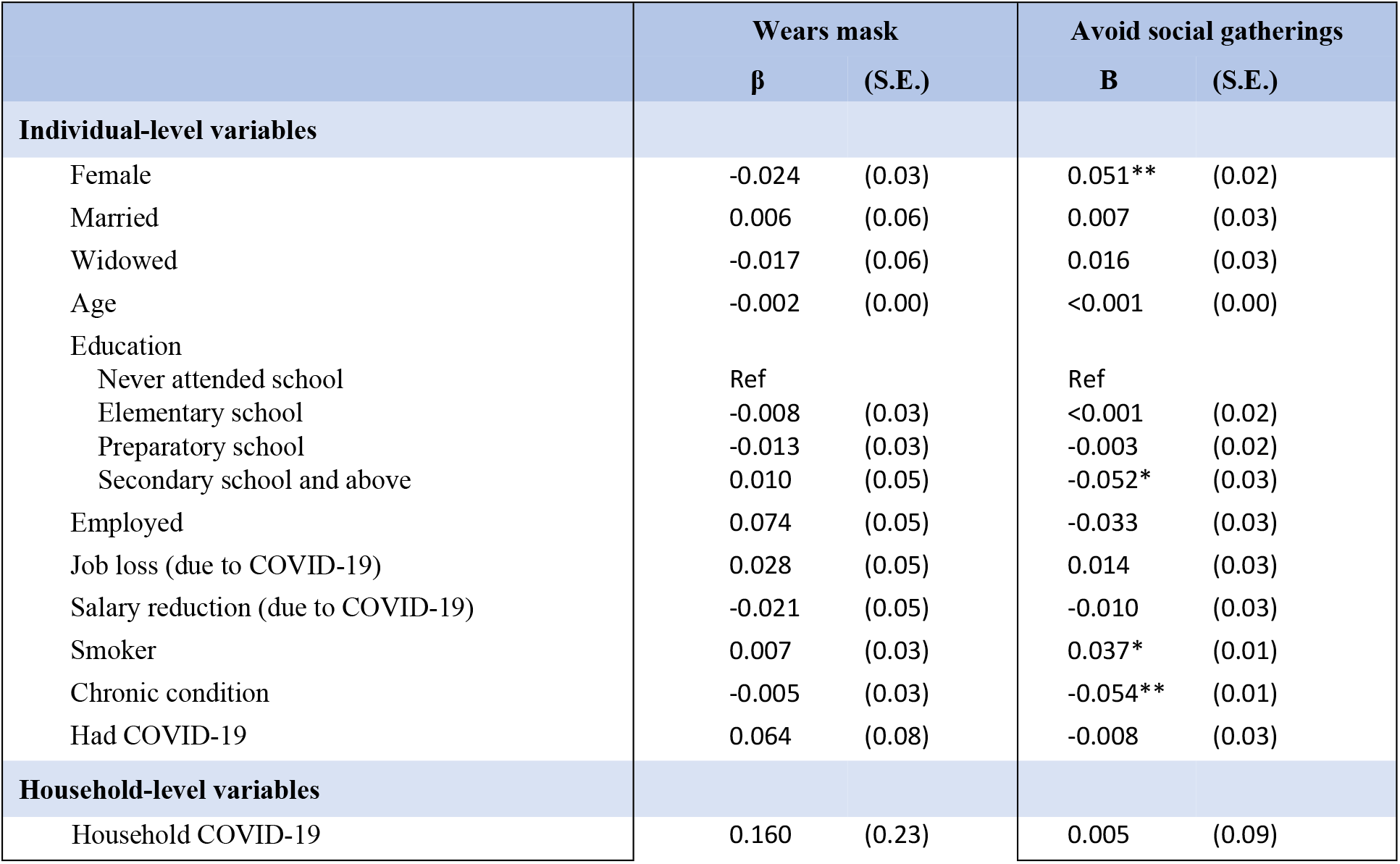
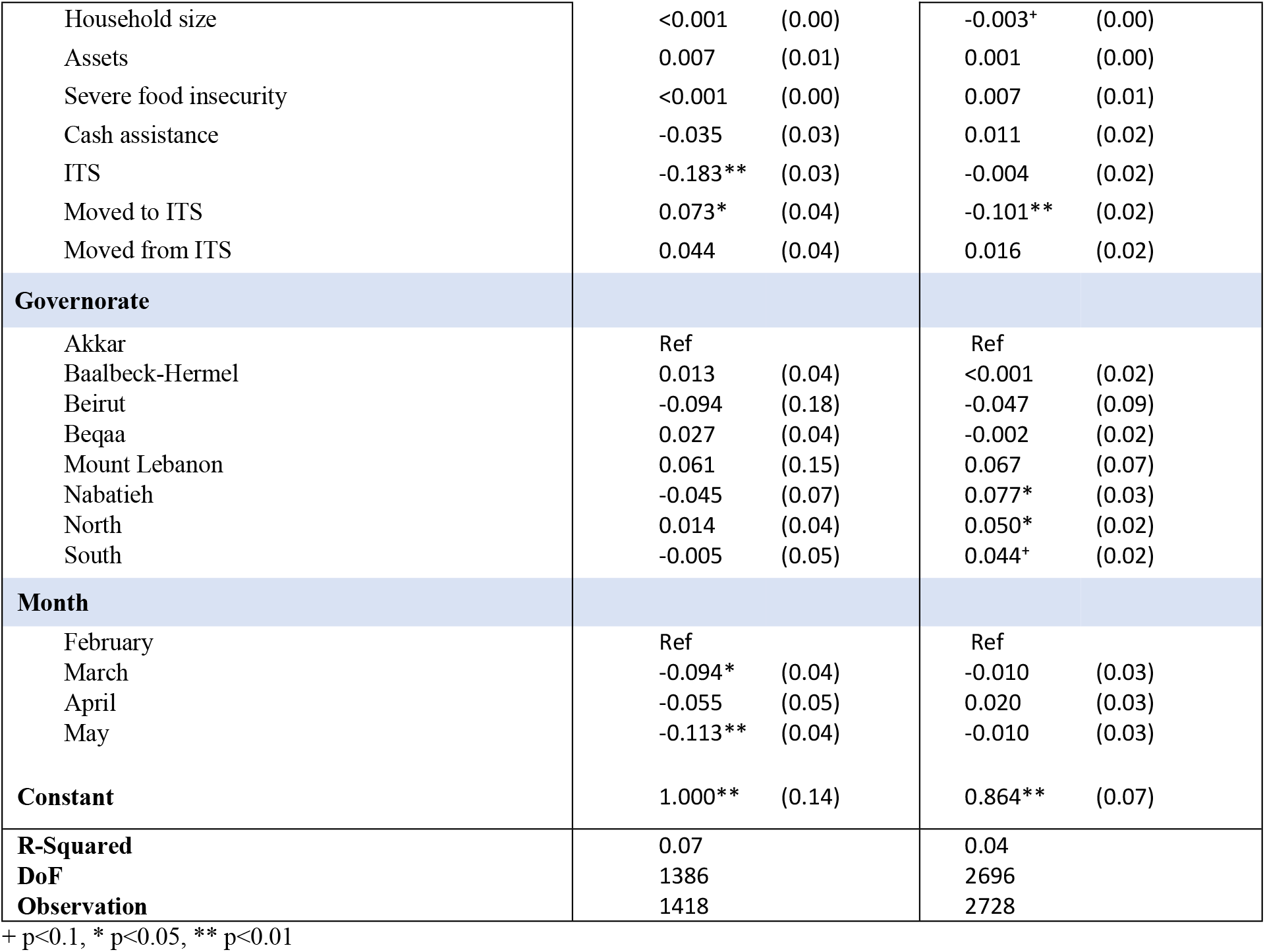
Sustained adherence after 6 months.

|  | Wears mask |  | Avoid social gatherings |  |
| --- | --- | --- | --- | --- |
| | $\beta$ | (S.E.) | B | (S.E.) |
| **Individual-level variables** |  |  |  |  |
| Female | -0.024 | (0.03) | 0.051** | (0.02) |
| Married | 0.006 | (0.06) | 0.007 | (0.03) |
| Widowed | -0.017 | (0.06) | 0.016 | (0.03) |
| Age | -0.002 | (0.00) | <0.001 | (0.00) |
| Education |  |  |  |  |
| Never attended school | Ref |  | Ref |  |
| Elementary school | -0.008 | (0.03) | <0.001 | (0.02) |
| Preparatory school | -0.013 | (0.03) | -0.003 | (0.02) |
| Secondary school and above | 0.010 | (0.05) | -0.052* | (0.03) |
| Employed | 0.074 | (0.05) | -0.033 | (0.03) |
| Job loss (due to COVID-19) | 0.028 | (0.05) | 0.014 | (0.03) |
| Salary reduction (due to COVID-19) | -0.021 | (0.05) | -0.010 | (0.03) |
| Smoker | 0.007 | (0.03) | 0.037* | (0.01) |
| Chronic condition | -0.005 | (0.03) | -0.054** | (0.01) |
| Had COVID-19 | 0.064 | (0.08) | -0.008 | (0.03) |
| **Household-level variables** |  |  |  |  |
| Household COVID-19 | 0.160 | (0.23) | 0.005 | (0.09) |
| Household size | <0.001 | (0.00) | -0.003<sup>+</sup> | (0.00) |
| Assets | 0.007 | (0.01) | 0.001 | (0.00) |
| Severe food insecurity | <0.001 | (0.00) | 0.007 | (0.01) |
| Cash assistance | -0.035 | (0.03) | 0.011 | (0.02) |
| ITS | -0.183** | (0.03) | -0.004 | (0.02) |
| Moved to ITS | 0.073* | (0.04) | -0.101** | (0.02) |
| Moved from ITS | 0.044 | (0.04) | 0.016 | (0.02) |
| **Governorate** |  |  |  |  |
| Akkar | Ref |  | Ref |  |
| Baalbeck-Hermel | 0.013 | (0.04) | <0.001 | (0.02) |
| Beirut | -0.094 | (0.18) | -0.047 | (0.09) |
| Beqaa | 0.027 | (0.04) | -0.002 | (0.02) |
| Mount Lebanon | 0.061 | (0.15) | 0.067 | (0.07) |
| Nabatieh | -0.045 | (0.07) | 0.077* | (0.03) |
| North | 0.014 | (0.04) | 0.050* | (0.02) |
| South | -0.005 | (0.05) | 0.044<sup>+</sup> | (0.02) |
| **Month** |  |  |  |  |
| February | Ref |  | Ref |  |
| March | -0.094* | (0.04) | -0.010 | (0.03) |
| April | -0.055 | (0.05) | 0.020 | (0.03) |
| May | -0.113** | (0.04) | -0.010 | (0.03) |
| **Constant** | 1.000** | (0.14) | 0.864** | (0.07) |
| **R-Squared** | 0.07 |  | 0.04 |  |
| **DoF** | 1386 |  | 2696 |  |
| **Observation** | 1418 |  | 2728 |  |

+ p<0.1, \* p<0.05, \*\* p<0.01

There is no significant distinction by gender, marital status, age, or education level between respondents who maintain the practice of wearing a mask and those who relent on it. Similarly, labor market variables, including employment, or changes in employment due to COVID-19 have no significant association with sustained adherence to mask wearing. Health variables, including a self-reported chronic condition, a personal or household-level history of COVID-19 also have no significant association with the continued practice of wearing a mask.

Instead, the only characteristics that show a significant correlation with sustained masking are household level variables related to the type of residence: residents of ITSs who initially report wearing a mask are significantly less likely to continue masking 6 months later, with a probability of sustained adherence 18.3 percentage points lower than for adherents. Interestingly, this effect is slightly attenuated (by 7.3 percentage points) for adherents who moved into an ITS during the 6 months between the two waves of data collection. This contrast in sustained adherence between original residents of ITSs and refugees who moved into ITSs suggests there may be pervasive community-based norms that affect individual level behavior. Other household level characteristics, including the household’s asset ownership, its food security status, its receipt of cash assistance (even COVID-19 related cash assistance) show no significant association with a respondent maintaining mask wearing.

While there are no geographic patterns in sustained adherence to masking, there is a noted and significant waning of adherence over time, as respondents who are surveyed later (March, April, May) are significantly less likely to sustain the practice of preventive mask wearing than respondents surveyed in February. This is also likely correlated with the timing of the peak of the surge of COVID-19 cases in early 2020 and the related lockdowns in January and February, which start to be eased in March.

Female respondents are significantly more likely to sustain abstinence from social gatherings, with the probability of maintaining the practice for 6 months 5.1 percentage points higher for women than men. Respondents with secondary or higher education levels on the other hand are more likely to have relented on the practice, conditional on having taken it up. Continued adherence does not appear to be significantly different for different ages or by marital status. Respondents who report having a chronic condition are a significant 5.4 percentage points more likely to give up on adherence, whereas smokers are 3.7 percentage points more likely to maintain adherence. Larger households are associated with a marginally significant and slightly lower likelihood of sustained adherence (by 1.2 percentage points for every 4 additional household members).

While we find some geographical differences in sustained adherence, with the North, South and Nabatieh governorates all associated with significantly higher likelihood of continuous compliance than Akkar, but respondents from households who moved into ITS in the 6 months between the two waves of observation are significantly less likely to maintain avoidance of social gatherings, with a 10.1 percentage point reduction in the likelihood of maintaining adherence. It is likely that moving into a densely populated and relatively well-defined community involves trying to integrate and socializing more easily.

Unlike in the case of maintaining masking, we find no time trend in the likelihood of sustaining the avoidance of social gatherings. Even though the strict lockdown of early 2021 started to be eased by mid-February, there is no significant difference in the rate of sustained adherence over six months between February and May.

## V. Robustness checks

The first four columns of Table 5 show the results from running the adherence regressions using a probit model, assuming normally distributed disturbances. The coefficients reported have been transformed to show marginal effects, calculated at the sample mean. While magnitudes are slightly different from the magnitudes estimated using the linear probability model in Table 3, the results are qualitatively very similar to those reported in the initial regressions. There is no change to note in the significance of any of the correlates of mask wearing or of avoiding social gatherings that were identified in the linear probability model.

**Table 5:**
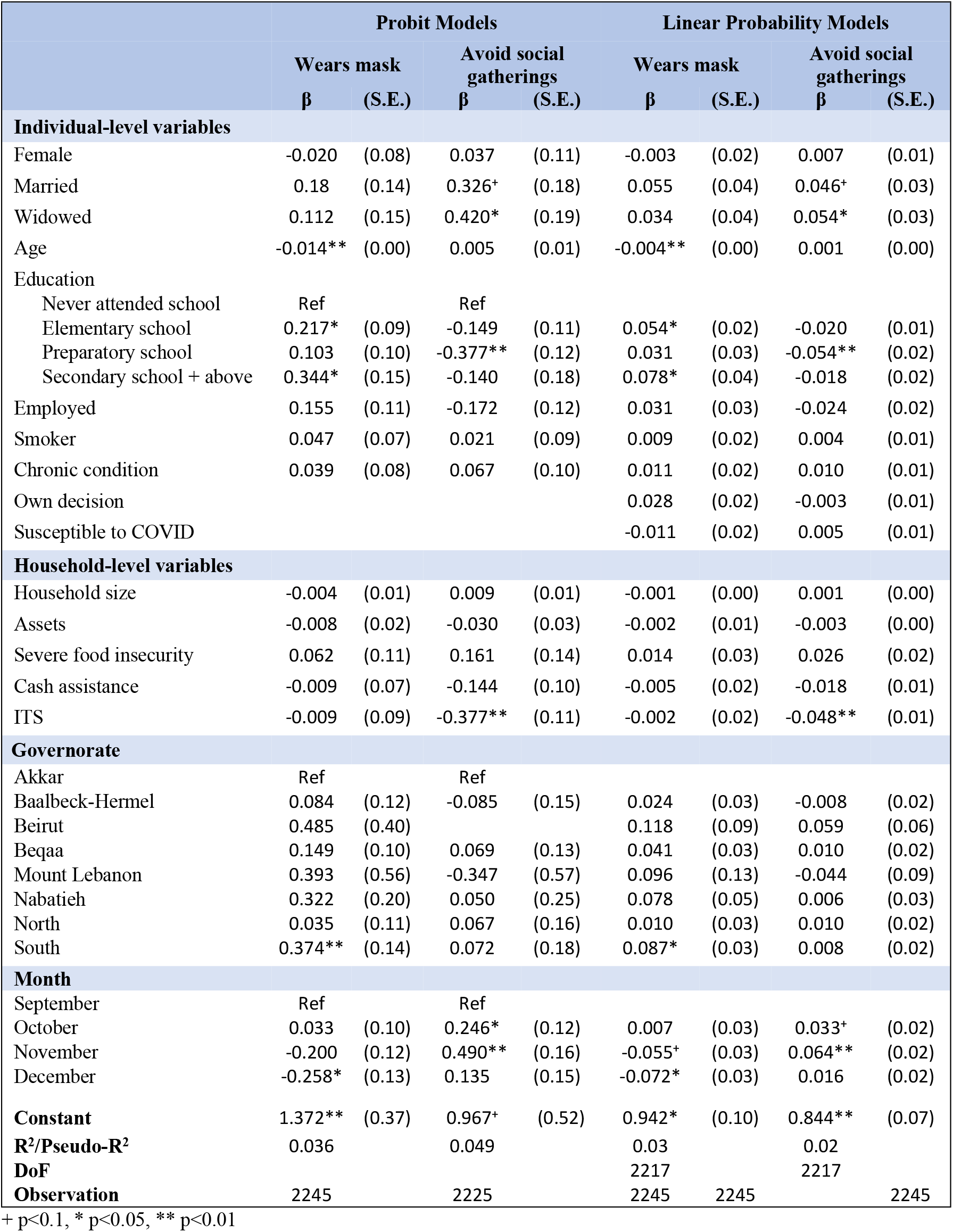
adherence to preventive measures.

Because adherence behavior tends to be highly idiosyncratic and personalized, the last four columns show the results from re-estimate the linear probability adherence regressions with two additional individual-level variables. The first measures self-reported susceptibility to COVID-19 (observed in the first survey) and the second indicates whether adherence to preventive behavior is based on the respondent’s own decision or on that of anyone else’s in the household. Neither of these variables is a significant correlate of adherence to preventive behavior, nor do they alter the results shown in Table 3 in any meaningful way.

Table 6 reports the results from similar checks for the robustness of the findings from Table 4. The estimates from running a probit model of continued adherence to preventive measures are qualitatively very similar to the estimates from the linear probability regression in Table 4. There is little change in the significance of the correlates, and while the magnitudes of the coefficients are evidently not identical to those of the linear probability model, assuming normally distributed errors does not alter the main takeaways from Table 4. The probit regression for continued adherence to mask wearing shows a marginally significant coefficient on the indicator of employment, with initial adherents who are employed 30.6 percentage points more likely to keep wearing masks (significant at the 10% level).

**Table 6:**
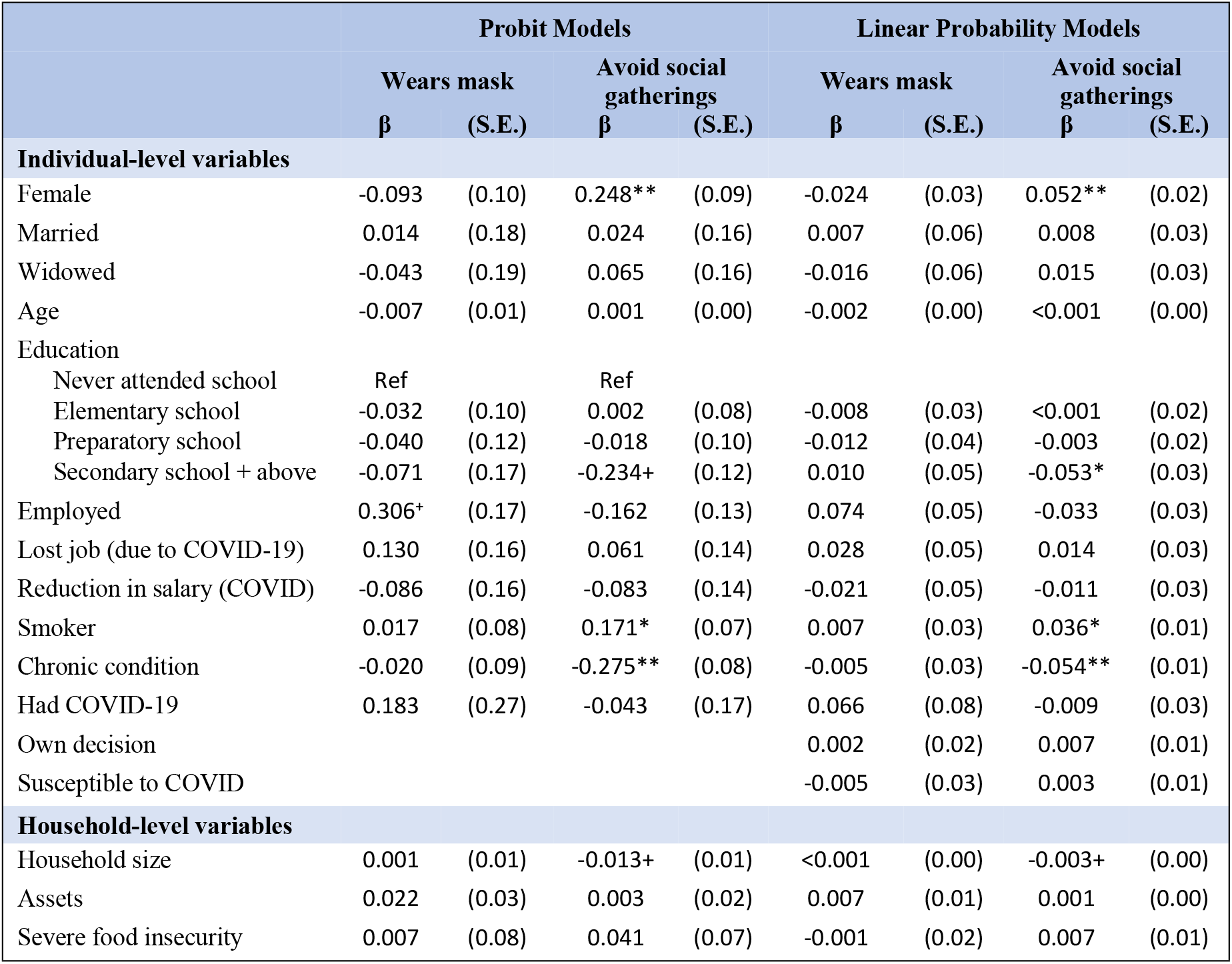
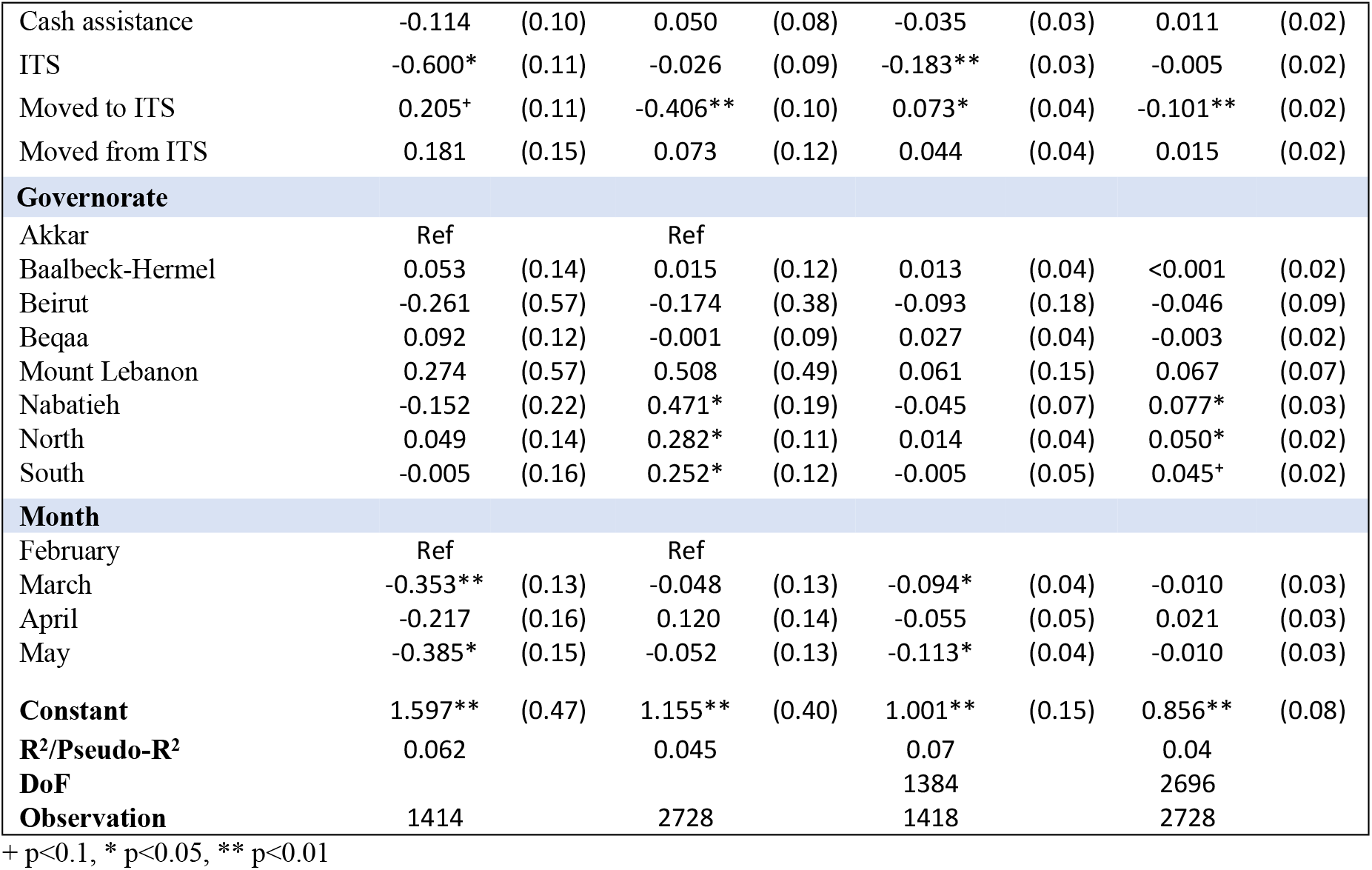
sustained adherence to preventive measures.

|  | Probit Models |  |  |  | Linear Probability Models |  |  |  |
| --- | --- | --- | --- | --- | --- | --- | --- | --- |
|  | Wears mask |  | Avoid social gatherings |  | Wears mask |  | Avoid social gatherings |  |
| | $\beta$ | (S.E.) | $\beta$ | (S.E.) | $\beta$ | (S.E.) | $\beta$ | (S.E.) |
| **Individual-level variables** |  |  |  |  |  |  |  |  |
| Female | -0.093 | (0.10) | 0.248** | (0.09) | -0.024 | (0.03) | 0.052** | (0.02) |
| Married | 0.014 | (0.18) | 0.024 | (0.16) | 0.007 | (0.06) | 0.008 | (0.03) |
| Widowed | -0.043 | (0.19) | 0.065 | (0.16) | -0.016 | (0.06) | 0.015 | (0.03) |
| Age | -0.007 | (0.01) | 0.001 | (0.00) | -0.002 | (0.00) | <0.001 | (0.00) |
| Education |  |  |  |  |  |  |  |  |
| Never attended school | Ref |  | Ref |  |  |  |  |  |
| Elementary school | -0.032 | (0.10) | 0.002 | (0.08) | -0.008 | (0.03) | <0.001 | (0.02) |
| Preparatory school | -0.040 | (0.12) | -0.018 | (0.10) | -0.012 | (0.04) | -0.003 | (0.02) |
| Secondary school + above | -0.071 | (0.17) | -0.234+ | (0.12) | 0.010 | (0.05) | -0.053* | (0.03) |
| Employed | 0.306+ | (0.17) | -0.162 | (0.13) | 0.074 | (0.05) | -0.033 | (0.03) |
| Lost job (due to COVID-19) | 0.130 | (0.16) | 0.061 | (0.14) | 0.028 | (0.05) | 0.014 | (0.03) |
| Reduction in salary (COVID) | -0.086 | (0.16) | -0.083 | (0.14) | -0.021 | (0.05) | -0.011 | (0.03) |
| Smoker | 0.017 | (0.08) | 0.171* | (0.07) | 0.007 | (0.03) | 0.036* | (0.01) |
| Chronic condition | -0.020 | (0.09) | -0.275** | (0.08) | -0.005 | (0.03) | -0.054** | (0.01) |
| Had COVID-19 | 0.183 | (0.27) | -0.043 | (0.17) | 0.066 | (0.08) | -0.009 | (0.03) |
| Own decision |  |  |  |  | 0.002 | (0.02) | 0.007 | (0.01) |
| Susceptible to COVID |  |  |  |  | -0.005 | (0.03) | 0.003 | (0.01) |
| **Household-level variables** |  |  |  |  |  |  |  |  |
| Household size | 0.001 | (0.01) | -0.013+ | (0.01) | <0.001 | (0.00) | -0.003+ | (0.00) |
| Assets | 0.022 | (0.03) | 0.003 | (0.02) | 0.007 | (0.01) | 0.001 | (0.00) |
| Severe food insecurity | 0.007 | (0.08) | 0.041 | (0.07) | -0.001 | (0.02) | 0.007 | (0.01) |
| Cash assistance | -0.114 | (0.10) | 0.050 | (0.08) | -0.035 | (0.03) | 0.011 | (0.02) |
| ITS | -0.600* | (0.11) | -0.026 | (0.09) | -0.183** | (0.03) | -0.005 | (0.02) |
| Moved to ITS | 0.205+ | (0.11) | -0.406** | (0.10) | 0.073* | (0.04) | -0.101** | (0.02) |
| Moved from ITS | 0.181 | (0.15) | 0.073 | (0.12) | 0.044 | (0.04) | 0.015 | (0.02) |
| **Governorate** |  |  |  |  |  |  |  |  |
| Akkar | Ref |  | Ref |  |  |  |  |  |
| Baalbeck-Hermel | 0.053 | (0.14) | 0.015 | (0.12) | 0.013 | (0.04) | <0.001 | (0.02) |
| Beirut | -0.261 | (0.57) | -0.174 | (0.38) | -0.093 | (0.18) | -0.046 | (0.09) |
| Beqaa | 0.092 | (0.12) | -0.001 | (0.09) | 0.027 | (0.04) | -0.003 | (0.02) |
| Mount Lebanon | 0.274 | (0.57) | 0.508 | (0.49) | 0.061 | (0.15) | 0.067 | (0.07) |
| Nabatieh | -0.152 | (0.22) | 0.471* | (0.19) | -0.045 | (0.07) | 0.077* | (0.03) |
| North | 0.049 | (0.14) | 0.282* | (0.11) | 0.014 | (0.04) | 0.050* | (0.02) |
| South | -0.005 | (0.16) | 0.252* | (0.12) | -0.005 | (0.05) | 0.045+ | (0.02) |
| **Month** |  |  |  |  |  |  |  |  |
| February | Ref |  | Ref |  |  |  |  |  |
| March | -0.353** | (0.13) | -0.048 | (0.13) | -0.094* | (0.04) | -0.010 | (0.03) |
| April | -0.217 | (0.16) | 0.120 | (0.14) | -0.055 | (0.05) | 0.021 | (0.03) |
| May | -0.385* | (0.15) | -0.052 | (0.13) | -0.113* | (0.04) | -0.010 | (0.03) |
| **Constant** | 1.597** | (0.47) | 1.155** | (0.40) | 1.001** | (0.15) | 0.856** | (0.08) |
| **R<sup>2</sup>/Pseudo-R<sup>2</sup>** | 0.062 |  | 0.045 |  | 0.07 |  | 0.04 |  |
| **DoF** |  |  |  |  | 1384 |  | 2696 |  |
| **Observation** | 1414 |  | 2728 |  | 1418 |  | 2728 |  |

+ p<0.1, \* p<0.05, \*\* p<0.01

The last columns of Table 6 add self-reported susceptibility to COVID-19 and a variable indicating whether adherence to preventive measures is a decision of the respondent themselves or of someone else in their household. These two variables turn out not to be significant correlates of continued adherence, and their inclusion has no effect worth noting on the coefficients of other variables.

Tables 5 and 6 suggest that the findings from the main regressions are neither driven by the linear functional form used in estimating the main regressions, nor are they sensitive to the inclusion of some of the other individual-level variables in the data that might conceivably be correlated to adherence behaviors.

## VI. Conclusion

Determining the correlates of adherence to preventive measures helps in identifying subgroups whose behavior has been riskier, which allows the design of interventions to target areas with a shortfall in prevention. Identifying variables associated with continued adherence helps update and refine interventions in the face of pandemic fatigue and changing disease dynamics.

It is clear that in this subpopulation of older Syrian refugees, targeting individuals with chronic diseases with information on COVID-19 prevention is needed to reverse the worrying finding that they appear to be more complacent about continued avoidance of social gatherings than those without chronic illness.

The lower continued adherence to mask wearing among residents of ITSs in particular points to factors beyond pandemic fatigue and that clearly should be taken into consideration in devising measures for disease control: the potential for community-based norms to determine individual-level behavior. This may be contributing to the waning in mask wearing, it may also be a contributor to the fact adherence to other preventive behavior (such as avoiding social gatherings) was significantly lower in ITSs, where lockdowns were less vigorously enforced and policed. Recognizing the pivotal effect of community-based norms in settings such as ITSs is essential in adapting current policy and designing future interventions.

The lack of an association with emergency COVID-19 cash assistance on the other hand indicates that refugees may prioritize other expenditures in the midst of the dual economic-health crisis. The possibility of expanding mask distribution to distribution sites outside of ITSs may be an important component of COVID-19 prevention efforts in this population.

## Data Availability

Data cannot be shared publicly because the data are on a vulnerable population (older Syrian refugees in Lebanon). Data are available from the Norwegian Refugee Council (NRC) Institutional Data Access/Ethics Committee (contact via Elsa Romera Moreno at NRC) for researchers who meet the criteria for access to confidential data.

## Footnotes

1 Asset ownership is a variable counting the number of affirmative answers to questions about the ownership of the following 15 assets: car, motorbike/scooter, van/pickup truck, bicycle, gas stove, oven, refrigerator, iron, heater, water heater, washing machine, TV, computer, mobile phone, access to the internet.

